# Study demands and health status among medical students in two German universities

**DOI:** 10.1101/2024.05.02.24306565

**Authors:** Amanda Voss, Susanne Dettmer, Mira Tschorn, Jan C. Zoellick

**Author notes:** Corresponding author: Jan C. Zoellick.

## Abstract

**Background and objectives:** Medical students regularly report high study demands and low mental health. We thus studied medical students in two different curricula in Germany investigating their study demands, study and life satisfaction, and overall and mental health with emphasis on contrasting beginners and advanced students.

**Design and methods:** We used online surveys with convenience samples through university mailing lists and student union channels. Uni1 participants (*N*=357; 70% female, *M*=24.83 years) were older than Uni2 participants (*N*=126; 79% female; *M*=23.39 years), but the cohorts did not differ further regarding sociodemographic variables. For analyses, we used *t*-tests for comparisons and correlations for associations.

**Results:** Students of both universities reported good mental and overall health as well as high satisfaction with their studies and lives, yet sleep difficulties were prevalent. Study demands were low to medium with the highest demands being learning activities (*M*=5.31; *SD*=2.19, scale 0-10), self-structuring (*M*=4.61; *SD*=2.01), and performance pressure (*M*=4.45; *SD*=2.27). The students in the reformed degree programme reported fewer issues with integrating theory and practice compared to those in the regular degree programme (*M*_*Uni1*_=3.38; *SD*_*Uni1*_=2.05; *M*_*Uni2*_=4.17; *SD*_*Uni2*_=2.06; *t*(430)=-3.53, *p*<.001).

**Discussion and conclusions:** Our sample was rather resilient regarding mental health and coping with study demands. The two universities showed little to no differences. With relative increased demands regarding learning activities, self-structuring, and performance pressure we suggest to focus on learning strategies for beginners and strategies for stress reduction to address, among other things, the reported sleep difficulties.

## 1. Introduction

Medical students regularly report excessive prevalence of depression [1], emotional exhaustion [2], burnout [3], and sleep difficulties [4] leading to low life satisfaction [5] as a result of stress. Impaired mental health is highly prevalent in university students with elevated levels of stress and depressive and anxiety symptoms [6].

Substance use as a malevolent coping mechanism is common in medical students [7] and translates into stress-burdened professional careers [8, 9]. In addition to these known factors, social support and structural conditions appear to be specific factors that can also impact the health of students [10, 11]. Understanding stress trajectories in their studies and creating an environment for better and healthier academic and professional achievements is one of the great challenges for medical faculties [12, 13]. Supporting resilience in students via specific intervention programmes has been proposed to mitigate stress trajectories [14-17]. Students burdened with poor mental health suffer disproportionately under high study demands. Furthermore, study demands seem to be a crucial source for stress [11]. An established measure for study demands in the German academic context differentiates six areas of demands: mode of science, learning activities, self-structuring, contact and cooperation, performance pressure and failure, and combining theory and practice [18]. These areas cover the broad range of expectations towards students ranging from organising oneself to performing in examinations or academic thinking and writing.

Students interact with the learning environment and perceive it in different ways based on their individual resources and characteristics [19]. Medical education in Germany is undergoing a dynamic reform process. After the introduction of the model clause in the Medical Licensing Regulations (ÄAppO) in 1999, numerous initiatives for the reform of medical studies have been implemented [20]. Currently, there are 13 reformed degree programs in Germany, including University 1 (Uni1), as well as 26 traditional degree programs, including University 2 (Uni2). A university curriculum can be understood as a learning environment that enables students to have learning experiences in order to achieve specific educational goals [21], and curricula can impact student satisfaction [22-24].

The purpose of this paper is to analyse medical students’ self-reported difficulties to cope with study demands, depression, exhaustion, sleep difficulties, general health status, and life as well as study satisfaction in two German universities, one structured following the reformed curriculum and one with a traditional curriculum. Exploratory studies have shown that these constructs are closely related to form stress trajectories – students with anxiety and stress tend to be overburdened by the workload leading to disengagement from their studies, depression, and burnout [25, 26]. The COVID-pandemic presented an additional burden potentially alleviating mental and overall health in students [27]. We suspect that the workload and anxiety is higher for beginners compared to advanced students. Since beginners have little experience in university exams, they lack a frame of reference for demands and realistic assessments of their own performance [28, 29]. Although previous research indicates no evidence for different rates of depressive symptoms between beginners compared to advanced students [30], this lack of a frame of reference might lead to inadequate expectations and additional stress.

### 1.1 Research questions

The following research questions are formulated: How high are the levels of study demands, depression, exhaustion, sleep difficulties, general health status, and life satisfaction in the whole sample? How do Uni1 and Uni2 students differ regarding their levels of study demands, depression, exhaustion, sleep difficulties, general health status, and life satisfaction? Are these constructs correlated? What are differences between cohorts (beginners and advanced students) regarding study demands and mental health outcomes?

## 2. Methods

### 2.1 Measures and definitions

We included the constructs depression, general health status, emotional exhaustion, sleep difficulties, life satisfaction, study satisfaction, and difficulties to cope with study demands in our survey. Table 1 defines the constructs and provides their measures. All constructs were transformed into POMP-scores [31] with a range 0 (low depression scores, bad health, few sleep difficulties, low life and study satisfaction, few difficulties in study demands) to 10 (high depression scores, good health, many sleep difficulties, high life and study satisfaction, many difficulties in study demands).

**Table 1.** Definition of constructs.

| Construct | Method | Range | Definition |
| --- | --- | --- | --- |
| Difficulties regarding study demands in... |  |  |  |
| ...mode of science | MWS [18] | 1 to 5 | Demands regarding the study content related to scientific language and thinking [18] |
| ...learning activities | MWS [18] | 1 to 5 | Personal demands linked to organising learning and studying [18] |
| ... self-structuring | MWS [18] | 1 to 5 | Organisational demands to structure contextual aspects of studying, e.g., assembling a study schedule [18] |
| ... contact and cooperation | MWS [18] | 1 to 5 | Social demands resulting from teamwork activities and forming bonds with other students [18] |
| ... performance pressure and failure | MWS [18] | 1 to 5 | Personal demands to cope with exam results or stress during exam preparation [18] |
| ... combining theory and practice | MWS [18] | 1 to 5 | Demands related to linking study contents to future occupation topics [18] |
| Depression | PHQ-2 [32] | 0 to 3 | "a period of at least 2 weeks during which there is either depressed mood or the loss of interest or pleasure in nearly all activities." [36] |
| Emotional exhaustion | OLBI [37] | 1 to 4 | "a consequence of intensive physical, affective, and cognitive strain, i. e., as a longterm consequence of prolonged exposure to certain job demands" [37] |
| Sleep difficulty | Self-devised | 1 to 5 | "Dyssomnias are primary disorders of initiating or maintaining sleep or of excessive sleepiness and are characterized by a disturbance in the amount, quality, or timing of sleep." [36] |
| Health status | Single item | 1 to 5 | Assessment of one's own health ("How is your health status generally?") |
| Life satisfaction | L-1 [38] | 0 to 100 | "cognitive process of assessing one's own wellbeing" [38] |
| Study satisfaction | Single item | 1 to 5 | Appraisal of students' study experience ("How satisfied are you overall with your study?") |

*Note.* The column range depicts the value range used in this study. After data collection, all constructs were transformed into POMP-scores [31] with a range 0 (few difficulties in study demands, low depression scores, low exhaustion, few sleep difficulties, bad health, and low life and study satisfaction,) to 10 (many difficulties in study demands, high depression scores, high exhaustion, many sleep difficulties, good health, and high life and study satisfaction).

### 2.2 Procedure

#### 2.2.1 Survey administration

We programmed an online questionnaire in German hosted on in-house servers of the respective university in accordance with European data protection laws. Participation was enabled in spring 2022. We invited students via the dean of studies mailing list and via the student union’s social media appearance.

#### 2.2.2 Ethical considerations

The ethics committee voted that no detailed examination of the pseudonymous study design was necessary (vote 21-393-ANF from 21 November 2021). Our study was carried out in accordance with the Declaration of Helsinki, including, but not limited to the anonymity of participants being guaranteed. Participants provided informed consent.

#### 2.2.3 Statistical analysis

Data were analysed using IBM SPSS Version 28. We calculated POMP scores for each variable in this analysis using the formula

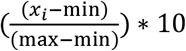

with *x*_*i*_ as individual observed values, *min* as the minimum value and *max* as the maximum value of the scale [31]. With their range between 0 and 10, POMP-scores can be intuitively interpreted in contrast to the varying original scores (e.g., 1 to 5 [18] or 0 to 3 [32]). The underlying linear transformation does not change the rank of values and is thus robust concerning parametric analyses, e.g. for *t*-tests, correlations, or Cronbach’s α [31]. We used descriptive statistics, *t*-tests, and correlations; results were considered statistically significant at *p* ≤ .05. For differences between beginners and advanced students, we created a dummy variable with values 0 (semesters 1-3) and 1 (semesters 8-10). We used pairwise deletion for missing data.

### 2.3 Sample characteristics

#### 2.3.1 Inclusion criteria

Medical students at Uni1 and Uni2 were invited to participate. We included participants who provided answers to ≥20% of the questions.

#### 2.3.2 Exclusion criteria

We excluded students of other subjects and other universities from participation. We also excluded datasets with answers to less than 20% of questions or incomplete datasets in cases of multiple participations.

#### 2.3.3 Sample size calculation

We did not perform a sample size calculation as this data collection was exploratory.

#### 2.3.4 Respondent characteristics

At Uni1, the welcome page was visited 656 times. 187 visitors were excluded immediately, because they provided no consent for participation or a subject of study other than medicine. We excluded 42 datasets for multiple participations. We additionally excluded 70 datasets with answers to <20% of questions. Thus, 357 medical students (70% female, *M* = 24.83 years, *SD* = 4.99 years) across all ten semesters of study (*Median* = 5^th^ semester) remained for analysis. At Uni1, 4,837 medical students were enrolled in the winter term 2021/22 resulting in a response rate of 7%.

At Uni2, the welcome page was visited 175 times. 33 visitors were excluded immediately, because they provided no consent for participation or a subject of study other than medicine. We excluded 4 datasets for multiple participations. We additionally excluded 12 datasets with answers to less than 20% of questions. Thus, 126 medical students (79% female, *M* = 24.41 years, *SD* = 3.47 years) across all ten semesters of study (*Median* = 5^th^ semester) remained for analysis. At Uni2, 2,828 medical students were enrolled in the winter term 2021/22 resulting in a response rate of 4%. Table 2 shows sample characteristics with tests for differences between universities.

**Table 2.** Sample characteristics

| | Uni1 | Uni2 | t / $\chi^2$ | p | d |
| --- | --- | --- | --- | --- | --- |
| N | 357 | 126 |  |  |  |
| Age (M, SD) | 24.83 (4.99) | 23.39 (3.47) | 3.53 | .000 | 0.31 |
| Gender (% women) | 71% | 79% | 3.98 | .137 | - |
| Semester (M, SD) | 5.31 (3.13) | 4.98 (2.62) | 1.15 | .252 | - |

*Note.* *M* = mean; *SD* = standard deviation; *t* = t-value for age and semester; $\chi^2$ = Chi-square value for gender; *p* = p-value test statistics; *d* = Cohen's *d*

## 3. Results

Our first research question aimed to assess mental and overall health as well as study demands in medical students. Students generally reported being mentally and overall healthy. They also reported being satisfied with their lives and their studies. Despite low mean depression scores (*M* = 3.16; *SD* = 2.75; range 0-10), 111 students (26% of valid responses) reported depression values above the clinically relevant threshold. Similarly, mean sleeping difficulty was low (*M* = 3.95; *SD* = 2.55), but only 161 participants (38% of valid responses) *never* or *rarely* experienced difficulties at all; 115 participants (27% of valid responses) reported *either* difficulties to fall asleep *or* sleep through the night; and the remaining 148 participants (35% of valid responses) reported difficulties of falling asleep *and* sleep through the night. Thus, reduced sleep quality was reported at least sometimes by the majority of the sample. Students also reported feeling medium exhausted in their studies. Table 3 shows descriptive statistics of the constructs. Reliability coefficients for all constructs were satisfying with the exception of self-structuring (Cronbach’s α = .49).

**Table 3.** Descriptive results of mental and overall health, study satisfaction, and study demands

| Construct | Range | N | *M* | *SD* | $\alpha$ | *t* | *p* |
| --- | --- | --- | --- | --- | --- | --- | --- |
| Mode of science | 0-10 | 318<sup>a</sup><br>113<sup>b</sup> | 4.06<br>3.87 | 2.26<br>2.33 | .78 | 0.78 | .437 |
| Learning activities | 0-10 | 319<sup>a</sup><br>113<sup>b</sup> | 5.28<br>5.39 | 2.18<br>2.21 | .78 | -0.44 | .660 |
| Self-structuring | 0-10 | 319<sup>a</sup><br>113<sup>b</sup> | 4.67<br>4.42 | 2.06<br>1.86 | .49 | 1.14 | .254 |
| Contact and cooperation | 0-10 | 319<sup>a</sup><br>113<sup>b</sup> | 4.36<br>4.02 | 2.10<br>2.17 | .69 | 1.47 | .143 |
| Performance pressure and failure | 0-10 | 319<sup>a</sup><br>113<sup>b</sup> | 4.35<br>4.73 | 2.26<br>2.26 | .74 | -1.51 | .132 |
| Combining theory and practice | 0-10 | 319<sup>a</sup><br>113<sup>b</sup> | 3.38<br>4.17 | 2.05<br>2.06 | .70 | -3.53 | .000 |
| Depression | 0-10 | 312<sup>a</sup><br>113<sup>b</sup> | 3.19<br>3.10 | 2.82<br>2.53 | .87 | 0.30 | .761 |
| Emotional exhaustion | 0-10 | 315<sup>a</sup><br>114<sup>b</sup> | 5.35<br>5.75 | 2.03<br>1.84 | .87 | -1.84 | .055 |
| Sleep difficulty | 0-10 | 312<sup>a</sup><br>114 | 3.91<br>4.06 | 2.57<br>2.49 | .69 | -0.54 | .589 |
| Health status | 0-10 | 317<sup>a</sup><br>113<sup>b</sup> | 7.33<br>7.48 | 2.34<br>1.96 | - | -0.63 | .527 |
| Life satisfaction | 0-10 | 319<sup>a</sup><br>113<sup>b</sup> | 6.58<br>6.55 | 2.28<br>2.11 | - | 0.11 | .910 |
| Study satisfaction | 0-10 | 331<br>115 | 6.75<br>6.50 | 2.37<br>2.29 | - | 0.99 | .332 |

*Note.* *M* = mean; *SD* = standard deviation; $\alpha$ = internal consistency (Cronbach's alpha); *t* = t-value for difference between Uni1 and Uni2 students; *p* = p-value for t-tests; <sup>a</sup> = Uni1 students; <sup>b</sup> = Uni2 students; Effect size for combining theory and practice: *d* = 0.39. Health status, life satisfaction, and study satisfaction were each measured with a single item, so Cronbach's $\alpha$ could not be calculated.

Our second exploratory question aimed at differences between Uni1 and Uni2. Uni2 students reported higher exhaustion that Uni1 students just short of statistical significance (*p* = .055). Students reported low to medium study demands with 3.38 ≤ *M* ≤ 5.39 on a scale ranging between 0 and 10. The most challenging aspects for both student groups were self-structuring, learning activities, and performance pressure, however in different orders. Significant differences were found in combining theory and practice which was more difficult for Uni2 students than for Uni1 students.

Our third exploratory question addressed correlations of constructs. All constructs regarding mental and overall health were closely related (.33 ≤ |*r*| ≤ .67; *p* < .01) indicating that respondents with low overall health also reported worse mental health, more exhaustion, and more sleep difficulties along with lower life and study satisfaction. All constructs regarding study demands were closely related (.24 ≤ |*r*| ≤ .61, *p* < .01), indicating that some students do well generally in the academic field while others struggle with learning, organising, and the scientific mode simultaneously. Communication and cooperation with fellow students showed the lowest correlations with other study demands. Comparing health and study constructs, mode of science was least associated with mental and overall health. Emotional exhaustion showed highest correlations with difficulties to cope with study demands. Study satisfaction was more closely related to practical application and study organisation than to contact with peers. Table 4 shows correlations between the constructs.

**Table 4.** Correlations between study demands, mental and overall health, and study satisfaction.

|  |  | 1 | 2 | 3 | 4 | 5 | 6 | 7 | 8 | 9 | 10 | 11 | 12 |
| --- | --- | --- | --- | --- | --- | --- | --- | --- | --- | --- | --- | --- | --- |
| 1 | Mode of science | - |  |  |  |  |  |  |  |  |  |  |  |
| 2 | Learning activities | .37** | - |  |  |  |  |  |  |  |  |  |  |
| 3 | Self-structuring | .35** | .49** | - |  |  |  |  |  |  |  |  |  |
| 4 | Contact and cooperation | .26** | .35** | .34** | - |  |  |  |  |  |  |  |  |
| 5 | Performance pressure and failure | .39** | .61** | .45** | .27** | - |  |  |  |  |  |  |  |
| 6 | Combining theory and practice | .31** | .38** | .37** | .24** | .35** | - |  |  |  |  |  |  |
| 7 | Depression | -.12* | .40** | .30** | .30** | .35** | .31** | - |  |  |  |  |  |
| 8 | Emotional exhaustion | .29** | .65** | .46** | .28** | .54** | .43** | .55** | - |  |  |  |  |
| 9 | Sleep difficulty | .11* | .30** | .21** | .20** | .29** | .16** | .40** | .41** | - |  |  |  |
| 10 | Health status | -.12* | -.30** | -.19** | -.23** | -.30** | -.17** | -.46** | -.44** | -.41** | - |  |  |
| 11 | Life satisfaction | -.18** | -.41** | -.33** | -.30** | -.30** | -.34* | -.67** | -.50** | -.33** | .52** | - |  |
| 12 | Study satisfaction | -.21** | -.34** | -.40** | -.21** | -.29** | -.48** | -.32** | -.44** | -.24** | .32** | .51** | - |

Note. $420 \leq N \leq 432$ ; \* $p < .05$ ; \*\* $p < .01$

Lastly, our fourth research question addressed differences in academic progression. Advanced medical students reported fewer difficulties to organise their learning and they suffered less from performance pressure than beginners – but only for Uni1 students. No differences were observed for Uni2 students. The other study demands did not differ between beginners and advanced students. Differences between beginners and advanced students of medicine are shown in Table 5.

**Table 5.** Differences between universities and between beginners and advanced students

| Construct | University | *N* | *M* | *SD* | *t* | *p* | *d* |
| --- | --- | --- | --- | --- | --- | --- | --- |
| Mode of science | Uni1 | 108<sup>a</sup> | 3.94 | 2.03 | -0.75 | .457 | - |
|  |  | 94<sup>b</sup> | 4.17 | 2.40 |  |  |  |
|  | Uni2 | 34<sup>a</sup> | 3.76 | 2.46 | -1.67 | .102 | - |
|  |  | 22<sup>b</sup> | 4.81 | 2.02 |  |  |  |
| Learning activities | Uni1 | 108<sup>a</sup> | 5.63 | 2.02 | 2.66 | .008 | 0.38 |
|  |  | 94<sup>b</sup> | 4.83 | 2.24 |  |  |  |
|  | Uni2 | 34<sup>a</sup> | 5.48 | 2.19 | 0.09 | .930 | - |
|  |  | 22<sup>b</sup> | 5.42 | 2.12 |  |  |  |
| Self-structuring | Uni1 | 108<sup>a</sup> | 4.63 | 2.14 | -0.23 | .818 | - |
|  |  | 94<sup>b</sup> | 4.70 | 2.10 |  |  |  |
|  | Uni2 | 34<sup>a</sup> | 4.20 | 1.80 | -0.93 | .358 | - |
|  |  | 22<sup>b</sup> | 4.66 | 1.79 |  |  |  |
| Contact and cooperation | Uni1 | 108<sup>a</sup> | 4.12 | 2.03 | -0.89 | .377 | - |
|  |  | 94<sup>b</sup> | 4.40 | 2.35 |  |  |  |
|  | Uni2 | 34<sup>a</sup> | 4.48 | 2.31 | 0.56 | .581 | - |
|  |  | 22<sup>b</sup> | 4.13 | 2.39 |  |  |  |
| Performance pressure and failure | Uni1 | 108<sup>a</sup> | 4.74 | 2.26 | 2.85 | .005 | 0.40 |
|  |  | 94<sup>b</sup> | 3.82 | 2.35 |  |  |  |
|  | Uni2 | 34<sup>a</sup> | 4.64 | 2.56 | -0.08 | .468 | - |
|  |  | 22<sup>b</sup> | 4.70 | 2.14 |  |  |  |
| Combining theory and practice | Uni1 | 108<sup>a</sup> | 3.16 | 1.95 | -1.09 | .282 | - |
|  |  | 94<sup>b</sup> | 3.48 | 2.22 |  |  |  |
|  | Uni2 | 34<sup>a</sup> | 3.85 | 2.11 | -1.43 | .079 | - |
|  |  | 22<sup>b</sup> | 4.72 | 2.37 |  |  |  |

Note. $N_{\text{beginners}} = 142$ ; $N_{\text{advanced}} = 116$ ; $M$ = mean; $SD$ = standard deviation; $t$ = t-value for difference between beginners and advanced of medical studies; $p$ = p-value for t-tests; $d$ = Cohen's $d$ ; <sup>a</sup> = beginners; <sup>b</sup> = advanced.

## 4. Discussion

We studied medical students in two different universities in Germany regarding their overall and mental health, coping with study demands, and their study and life satisfaction.

Results revealed students of both universities reporting good overall and mental health as well as high satisfaction with their studies and lives. However, 26% students reported depressive symptoms above the clinically relevant cut-point. This rate was slightly lower than in a study investigating students at five German universities during the COVID-pandemic [33], but substantially higher to the general German population, where 10.3% (17-29-year-olds) and 10.9% (30-39-year-olds) reported elevated depressive symptoms measured with the PHQ-2 [34]. Sleep difficulties as a particular aspect of mental health were prevalent in the majority with 62% of respondents reporting sleep disturbances at least sometimes. This corresponds to 52.7% of 25,735 medical students reporting poor sleep quality that were included in a meta-analysis [4]. In comparison, 36% of the general German population reported poor sleep quality [35]. Thus, students reported seemingly conflicting findings of good overall health and simultaneously more sleep difficulties and depressive symptoms than the general population. However, sleep as a specific measure can be seen as an early indicator of mental health difficulties compared to manifest depressive symptoms or declined overall health. We thus suggest to intervene early to interrupt a pathway from current early indicators of stress and struggle towards chronified versions of stress and struggle.

Study demands most difficult to cope with were learning activities (*M* = 5.31), self-structuring (*M* = 4.61), and performance pressure (*M* = 4.45). These three demands were also the most difficult to cope with in the representative samples of Jänsch and Bosse [18] across degree programmes with somewhat lower POMP-scores (*M*_*LA*_ = 4.60; *M*_*SS*_ = 3.93; *M*_*PP*_ = 4.30). We observed significant differences between the two universities only regarding Uni1 students reporting lower difficulties to combine theory and practice than Uni2 students. For both universities, more difficulties combining theory and practice were relatively strongly correlated with depression (*r* = .31), emotional exhaustion (*r* = .43), and study satisfaction (*r* = -.48).

Differences between beginners and advanced students to cope with study demands were inconclusive. Advanced students had lower difficulties to cope with learning activities and performance pressure at Uni1, but not at Uni2 – all other study demands did not yield differences in study progression. These differences can be explained via experience – advanced students have built up a wealth of experience in exam situations and can better assess their expectations and competencies compared to beginners.

Surprisingly, academic progress did not correspond to better assessment of contact and cooperation between students at either university. First to third semester students in winter term 2021 had primarily experienced online teaching whereas their more experienced counterparts studied the majority of their academic career in-person. Yet, online teaching with fewer opportunities to get in contact with fellow students did not compromise the social embeddedness in beginners compared to advanced students.

With our findings, we did not replicate the results of previous studies presenting medical students as depressed, overworked, exhausted, and with low quality of life [1-3, 5]. However, sleep difficulties reported in the literature [4] were also prevalent in our sample. We thus suggest to develop and offer programmes for students supporting their mental health with particular focus on sleep.

Mental and overall health were correlated, as well as the constructs of study demands. This suggests that some students struggle particularly whereas others are able to cope with study demands more easily. This supports the call to identify vulnerable, struggling students to provide adequate, individual support in their studies.

### 4.1 Limitations

Convenience samples were used for both universities; we invited students to participate via e-mail lists and social media activities of the student unions. Thus, particularly students who are keen to provide feedback on their study experience were inclined to answer presenting selection bias. Despite convenience sampling, gender ratios, age, and median semester of studies were representative of the student body and did not differ between the two universities. We are thus confident that our findings are informative for the respective study populations.

## 5. Conclusions

Our sample was rather resilient regarding mental health and coping with study demands. Curricular influence on our outcomes seemed comparatively minor, which might result from increased online teaching during the pandemic. We suggest to focus on learning strategies for beginners and strategies for stress reduction to address, among other things, the reported sleep difficulties.

## Data Availability

All data produced in the present study are available upon reasonable request to the authors.

## Acknowledgements

The authors would like to thank Luise Chassol from the Uni1 student union for support in finalising the questionnaire and Wiebke Höppner from the Uni1 dean’s office for supporting student recruitment. The authors would also like to thank the Uni1 student assistants Anton Horeis and Janek Wilhelm for survey programming and data cleaning.

## Contributions statement

AV, SD, and JZ developed the study design and questionnaire, and collected the data; JZ analysed the data and created the first manuscript draft; all authors interpreted the data and substantively revised the manuscript; all authors read and approved the final manuscript.

